# ACE2 expression by colonic epithelial cells is associated with viral infection, immunity and energy metabolism

**DOI:** 10.1101/2020.02.05.20020545

**Authors:** Jun Wang, Shanmeizi Zhao, Ming Liu, Zhiyao Zhao, Yiping Xu, Ping Wang, Meng Lin, Yanhui Xu, Bing Huang, Xiaoyu Zuo, Zhanghua Chen, Fan Bai, Jun Cui, Andrew M Lew, Jincun Zhao, Yan Zhang, Hai-Bin Luo, Yuxia Zhang

**Author notes:** These authors contributed equally (Jun Wang, Shanmeizi Zhao).

## Abstract

Respiratory disease caused by the 2019 novel coronavirus (2019-nCoV) pneumonia first emerged in Wuhan, Hubei Province, China, in December 2019 and spread rapidly to other provinces and other countries. Angiotensin-converting enzyme 2 (ACE2) is the receptor for SARS-CoV and has been suggested to be also the receptor for 2019-nCoV. Paradoxically, ACE2 expression in the lung protects mice from SARS-CoV spike protein induced lung injury by attenuating the renin-angiotensin system. In the intestine, ACE2 also suppresses intestinal inflammation by maintaining amino acid homeostasis, antimicrobial peptide expression and ecology of the gut microbiome. Upon analysis of single cell-RNA sequencing data from control subjects and those with colitis or inflammatory bowel disease (IBD), we found that *ACE2* expression in the colonocytes was positively associated with genes regulating viral infection, innate and cellular immunity, but was negatively associated with viral transcription, protein translation, humoral immunity, phagocytosis and complement activation. In summary, we suggest that ACE2 may play dual roles in mediating the susceptibility and immunity of 2019-nCoV infection.

---

Patients infected by 2019-nCoV can develop acute respiratory distress syndrome (ARDS) and sepsis. However, there is mounting evidences that the gastrointestinal tract may be an alternative route for 2019-nCoV infection. One (3%) of the first 41 reported cases had diarrhea [1]. In another report describing a familial cluster, 2 of the 6 family members (33%) developed diarrhea[2]. Furthermore, positive 2019-nCoV was detected from the stools of a patient with loose of bowel movement in the United States [3].

There have been many studies on the proteins associated with host entry by the SARS-CoV. Cleavage of Spike protein by tissue factor Xa is required for the infectivity of SARS-CoV [4]. Rivaroxaban is an oral anticoagulant used to combat acute pulmonary embolism with or without deep-vein thrombosis [5]. As an Xa inhibitor, it may thus offer protection for 2019-nCoV pneumonia. Angiotensin I converting enzyme 2 (ACE2) is the SARS-CoV receptor [6]. It also has been linked to 2019-nCoV infection; ACE2 interacts with the Spike protein and mediates 2019-nCoV infection of the type II alveolar cells of the lung [7].

Paradoxically, although ACE2 mediates viral entry to the host, its deficiency worsens lung injury by activating the renin-angiotensin system (RAS) in experimental models [6]. The circulating RAS regulates blood pressure and fluid homeostasis. Local tissue-based RAS exacerbates pulmonary hypertension, acute lung injury and experimental lung fibrosis [8]. Thus, blocking of the Angiotensin II receptor type I (AT1R) was associated with reduced SARS-CoV spike protein mediated lung injury [6] and reduced pulmonary hypertension in experimental models [9]. The AT1R blocker valsartan has recently been reported to improve clinical outcomes for patients with chronic obstructive pulmonary disease (COPD) complicated with pulmonary hypertension (DOI:10.1183/13993003.congress-2019.PA2469). In the PARADIGM-HF trial, valsartan and the neprilysin inhibitor prodrug sacubitril (Valsartan/sacubitril, branded Entresto by Novartis) also significantly reduced mortality and hospitalization for patients with heart failure [10].

Apart from the lung, ACE2 is also associated with gut function. ACE2 expression in the gut epithelial cells is required for maintaining amino acid homeostasis, antimicrobial peptide expression and the ecology of gut microbiome in the intestine [11, 12]. We had previously generated a single-cell RNA sequencing dataset from control subjects and those with colitis or IBD [13]. Using this dataset, we examined ACE2 expression and found that it was specifically and highly expressed by the colonocytes (**Figure 1 A-B**). Compared with other epithelial subtypes, colonocytes overexpressed genes regulating viral entry, budding and releasing from the host cells (*VAPA, CHMP4C, CHMP1A, CHMP2B, CHMP4B, CHMP1B, VPS37B, PDCD6IP, VPS4B, CHMP3, VPS28, CHMP2A, PVRL2, CDH1*, and *MVB12A*). Genes involved in nucleic acid sensing and type I, type III interferon signaling (*R2RX4, DDX58, NLRC5, IFNAR2, IFNGR1* and *IFNGR2*), pro-inflammatory cytokine and chemokine (*IL18, IL32, CXCL16, CXCL3, CXCL2, CXCL1, CCL28, CCL20*) production, antigen presentation via MHC class I molecules (*HLA-A, HLA-B, HLA-C*, and *HLA-E)* and cytokine receptors (*IL2G, IL10RB*, and *IL6R*) were also enriched in colonocytes (**Figure 1C**).

**Figure 1.**
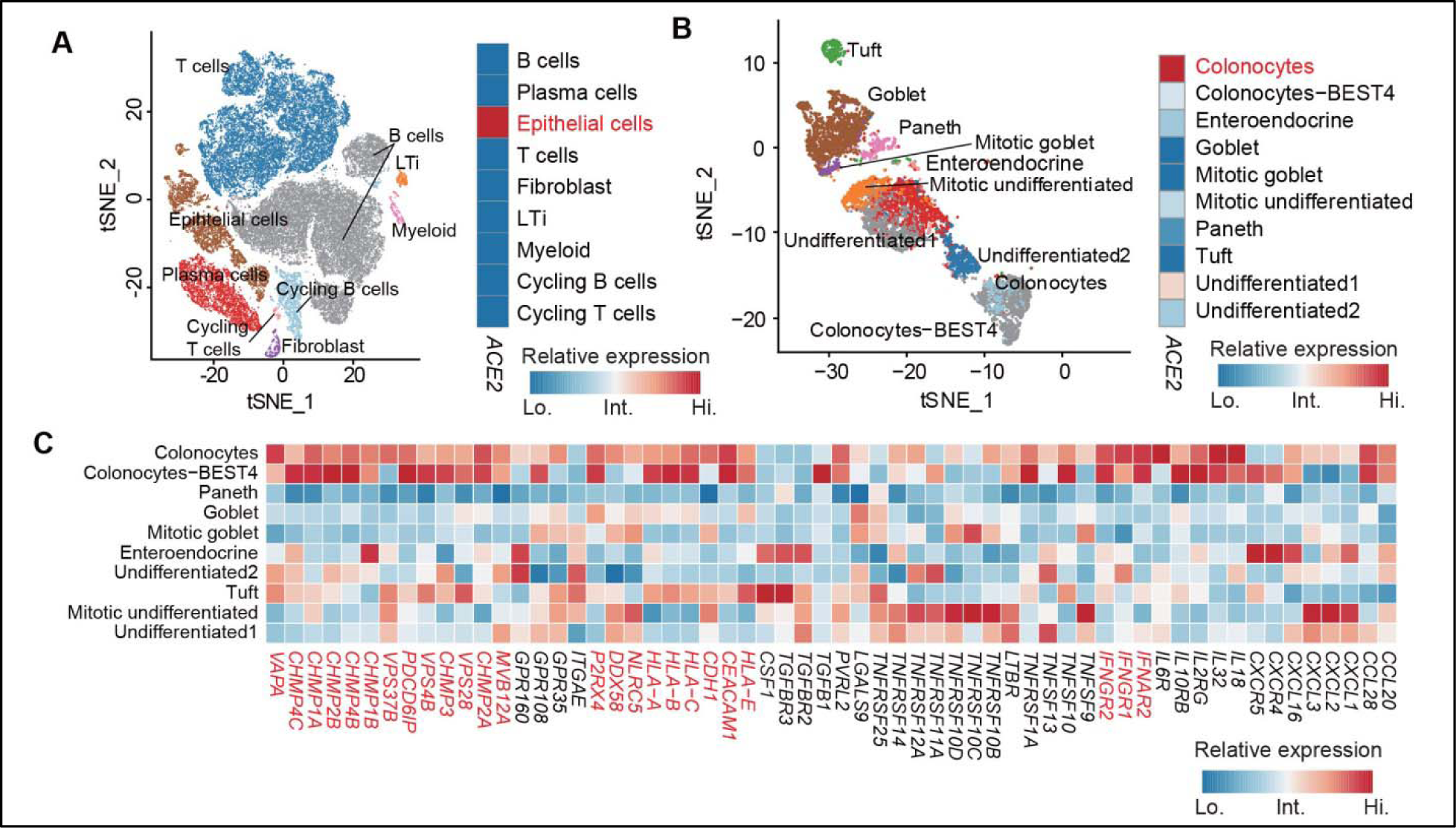
ACE2 was highly and specifically expressed by colonocytes. **A**. Left panel: two-dimensional tSNE plot displaying major cell clusters; right panel: heatmap displaying *ACE2* expression across major cell clusters. **B**. Left panel: distribution of 10 epithelial sub-clusters on the tSNE plot; right panel: heatmap showing *ACE2* expression across epithelial clusters. **C**. Heatmap plot showing expression of selected genes across 10 epithelial clusters.

Given that ACE2 plays dual roles in mediating viral entry and preventing tissue injury [6, 11, 12, 14], we analyzed genes that are co-expressed with ACE2 in the colonic epithelial cells. In total, 3420 and 2136 genes were positively and negatively correlated with *ACE2* expression, respectively (**Supplementary table 1 and Figure 2A**). In Gene Ontology (GO) analysis, genes positively correlated with *ACE2* expression were enriched in viral infection and egress (viral budding via host ESCRT complex, positive regulation of viral release from host cells, viral entry into host cells), innate immune responses (type I interferon signaling pathway and toll-like receptor signaling pathway), NK and T cell cytotoxicity (interferon-gamma-mediated signaling pathway, regulation of natural killer cell mediated immunity, positive regulation of T cell mediated cytotoxicity), energy metabolism (triglyceride biosynthetic process, pyruvate metabolic process, NADH metabolic process, mitochondrial electron transport), inflammation and apoptosis (positive regulation of I-kappaB kinase/NF-kappaB signaling and positive regulation of apoptotic signaling pathway). In contrast, genes negatively correlated with *ACE2* were enriched for humoral immunity (positive regulation of B cell activation, antibacterial humoral response, mature B cell differentiation, B cell receptor signaling pathway, immunoglobulin production), protein translation (ribosomal big and small subunit assembly and translational initiation), phagocytosis (Fc-epsilon receptor signaling pathway, Fc-gamma receptor signaling pathway involved in phagocytosis, phagocytosis recognition) and classical pathway of complement activation (**Figure 2B**).

**Figure 2.**
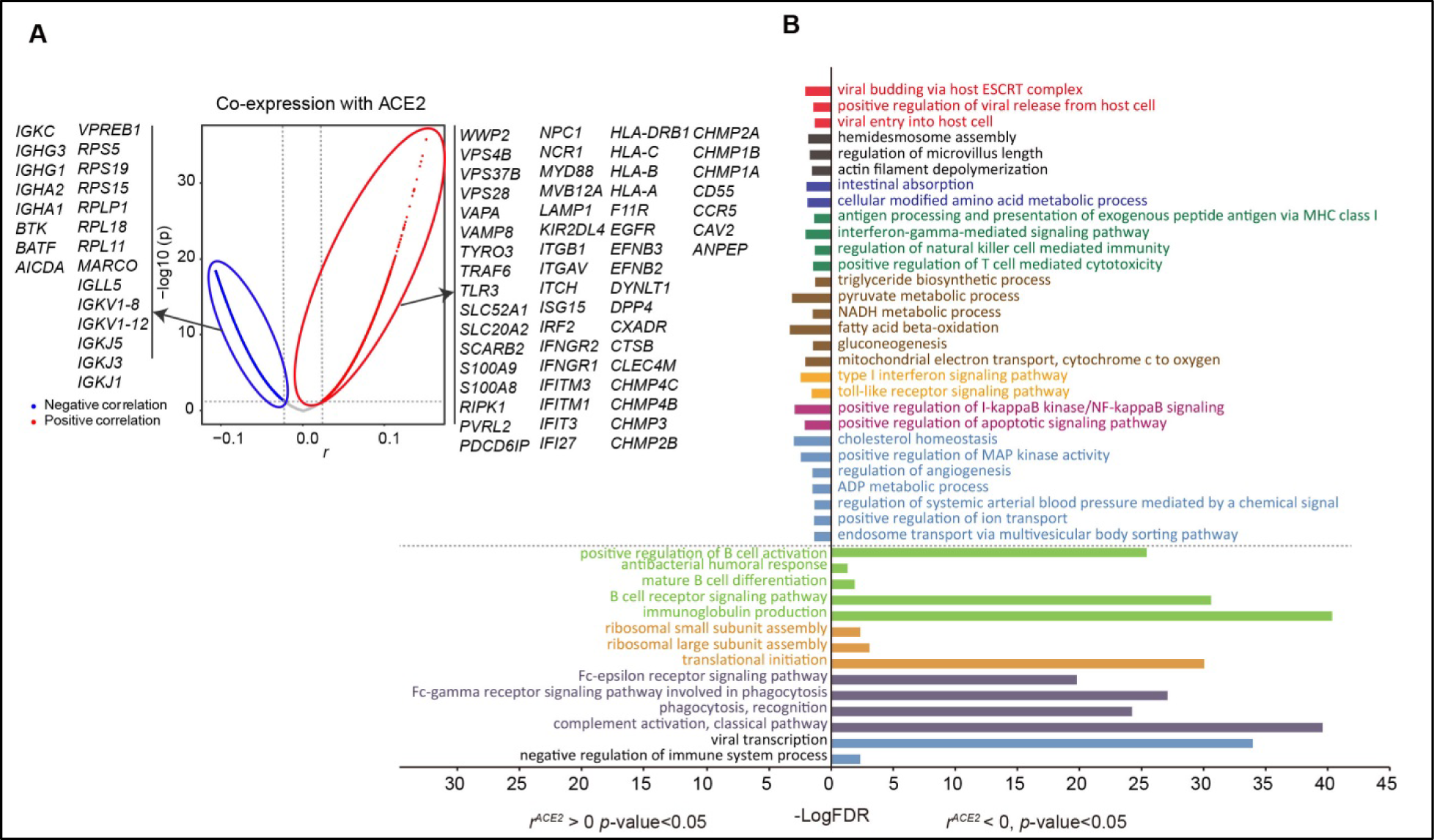
Genes correlated with *ACE2* expression in colonocytes. **A**. Valcano plot displaying genes co-expressed with *ACE2*. **B**. Bar plot presenting significantly enriched GO terms obtained from GO enrichment analysis performed with aforementioned *ACE2* co-expressed genes.

## Discussion

Here we report the expression pattern of ACE2 in the human intestine at single-cell resolution. Our data suggest that ACE2 is specifically expressed by colonocytes and positively associated with viral entry and egress, consistent with observations that the gastrointestinal tract is a potential route for 2019-nCoV infection. In supportive of the view that ACE2 expression prevents intestinal inflammation [11, 12], bioinformatic analysis suggested that genes regulating type I and type III innate immunity, NK and T cell mediated cytotoxicity and energy metabolism are positively associated with ACE2 expression. By contrast, humoral immunity, phagocytosis and complement activation are negatively associated with ACE2 expression. These findings may facilitate concurrent and future investigations of the nature and function of the immune system at systemic and local levels in patients with 2019-nCoV pneumonia.

Notably, SARS-CoV spike protein engagement downregulates ACE2 expression and activates the RAS [6]. Given that hypertension is common in severe 2019-nCoV pneumonia [1], it is highly likely that the RAS is activated in the lungs of patients with severe pneumonia. The Xa inhibitor Rivaroxaban [5] and the AT1R blocker valsartan (DOI:10.1183/13993003.congress-2019.PA2469) are effective in relieving pulmonary injury and embolism in experimental models and clinical trials, their efficacy in treating 2019-nCoV may worth consideration. Dipyridamole is another anti-coagulation drug. In a pilot trial, dipyridamole administration promoted mucosal healing and improved clinical symptoms in patients with colitis or IBD [13]. More importantly, published clinical studies [15-20], including those conducted in China (DOI: CNKI:SUN:XIYI.0.2010-12-026;[21]; CNKI:SUN:YXLT.0.2014-32-031), have demonstrated that dipyridamole has broad spectrum antiviral activity against virus infections. It may also be worth investigating the therapeutic effects of dipyridamole in patients infected by 2019-nCoV.

## Methods

### Ethical statement

The study procedures (ID: 2017021504) were approved by the Medical Ethics Committee of Guangzhou Women and Children’s Medical Center. The implementations were in concordance with the International Ethical Guidelines for Research Involving Human Subjects stated in the Helsinki Declaration. Informed written consents from the legal guardians of all participants were obtained before conducting these procedures.

### Single cell RNA sequencing data analysis

Colonic biopsies from 17 children (6 control, 6 colitis, 2 ulcerative colitis, 3 Crohn’s disease) were subjected to single-cell suspension preparation, 5 ‘ gene expression libraries construction, and 10x Genomics single-cell RNA sequencing [13]. Cellranger v2.1 pipeline (10x genomics) was utilized to demultiplex the cellular barcodes and align reads to the human transcriptome (build GRCh38). The unique molecular identifier (UMI) count matrix was converted to Seurat object using the R package Seurat v2.3.4 [22]. After quality control and data normalization, dimension reduction and cell clustering were performed [13]. Different cell types were identified by mapping canonical marker genes in the two-dimensional t-Distributed Stochastic Neighbor embedding (tSNE) map [13].

### Co-expression analysis

Normalized single-cell RNA expression data was used for co-expression analysis. Pearson’s correlation analysis was performed between each gene in the single-cell transcriptome and *ACE2*. Significantly correlated genes were chosen (p-value<0.05) for downstream GO enrichment analysis via GENEONTOLOGY at http://geneontology.org/. GO terms with False Detection Rate (FDR) <0.05 were selected.

## Data Availability

The single-cell RNAseq data was from our recently published work entitled "Profiling of Pediatric-Onset Colitis and IBD Reveals Common Pathogenics and Therapeutic Pathways"( Cell, 2019. 179(5): p. 1160-1176 e24.)

## Acknowledgments

National Natural Sc The ience Foundation of China (91742109, 31770978, 31722003, 31770925, 31370847, 81770552), National Key Research and Development Program (2016YFC0900102), National Science and Technology Major Project (2018ZX10302205), Guangdong Provincial Key Laboratory of Research in Structural Birth Defect Disease (2019B030301004), Guangzhou Women and Children’s Medical Center Fund (5001-3001032) and National Health and Medical Research Council of Australia (1037321, 1105209, 1143976, and 1080321 to A.L.) funded this study.

## Author Contributions

Yuxia.Z., H-B.L., and Yan.Z. conceived of and supervised the project. J.W. performed the bioinformatic analysis. S.Z., M.L., Z. Z., Yiping.X, P.W, M. L., Yanhui.X., B.H., Xiaoyu.Z. and Z.C. reviewed and discussed the manuscript. Yuxia.Z. and J.W. wrote the manuscript with significant input from A.M.L., J.C., F.B. and All authors discussed and approved the manuscript.

**Supplementary Table 1** Genes correlated with *ACE2* expression.

## Notes

### Competing Interest Statement

The authors have declared no competing interest.

### Funding Statement

The National Natural Science Foundation of China (91742109, 31770978, 31722003, 31770925, 31370847, 81770552), National Key Research and Development Program (2016YFC0900102), National Science and Technology Major Project (2018ZX10302205), Guangdong Provincial Key Laboratory of Research in Structural Birth Defect Disease (2019B030301004), Guangzhou Women and Children’s Medical Center Fund (5001-3001032) and National Health and Medical Research Council of Australia (1037321, 1105209, 1143976, and 1080321 to A.L.) funded this study.

